# Invasive Fungal Infection in Childhood Embryonal Brain Tumour Treatment: A 10-year Review

**DOI:** 10.64898/2026.08.13.26359732

**Authors:** Sean M Carter, Aniket Chawla, Martin Campbell, David D. Eisenstat, Heather Weerdenburg, Dong-Anh Khuong-Quang, Gabrielle M Haeusler

## Abstract

**Background:** Invasive fungal infection (IFI) is well recognised in children with acute leukaemia and allogeneic haematopoietic stem-cell transplantation but is poorly characterised in children with brain tumours. Children receiving intensive therapy for embryonal brain tumours (EBTs) have multiple potential risk exposures including corticosteroids, central venous access, neurosurgical devices, mucosal injury and myelosuppressive chemotherapy with, in selected protocols, autologous stem-cell rescue.

**Methods:** We performed a single-centre retrospective cohort study of children aged 0–18 years treated for EBTs between 2015-2025. IFIs were classified as proven, probable, possible, or modified possible using EORTC/MSGERC and TERIFIC criteria. Clinical characteristics, treatment exposures, timing, microbiology and outcomes were described. IFI prevalence was calculated using exact binomial confidence intervals. Exploratory Cox proportional hazards analyses assessed associations with clinical and treatment factors.

**Results:** Seventy-seven patients were included. Fourteen patients experienced 15 IFI episodes, giving a patient-level IFI prevalence of 18.2% (95% CI, 10.3– 28.6%). Proven or probable IFI occurred in seven patients (9.1%; 95% CI, 3.7–17.8%). Nine episodes had microbiological evidence. Non-mould pathogens predominated, accounting for six of nine identified pathogens. Treatment on ACNS0334/ACNS0333 was associated with a lower hazard of proven/probable IFI compared with SJMB12 (HR 0.062; 95% CI, 0.002–0.78; p=0.031). Two patients had chemotherapy delays exceeding one month, one had persistent infection at 12 months; no deaths were directly attributed to IFI. Three patients received antifungal prophylaxis.

**Conclusion:** Rates of IFI following intensive embryonal brain tumour therapy were comparable to those in other high-risk oncology populations. Local consideration of antifungal prophylaxis is warranted.

**Key Points:**

1. Among children receiving intensive therapy for embryonal brain tumours, 18.2% developed an IFI of any category and 9.1% developed proven or probable IFI.
2. Non-mould infections predominated, accounting for 6/9 mycologically identified episodes (67%).

**Importance of the Study:** Invasive fungal infection (IFI) risk is poorly described during paediatric brain tumour treatment. Incidence has not previously been reported in embryonal brain tumours despite these children having multiple risk exposures including corticosteroids, central venous access, neurosurgical devices, mucosal injury and myelosuppressive chemotherapy with, in selected protocols, autologous stem-cell rescue. Our study addresses this gap by describing IFI incidence, timing and microbiology during intensive EBT therapy, in a denominator-defined cohort. Our findings help identify EBT treatment as a potentially under-recognised setting for IFIs and support prospective evaluation of risk-stratified prevention strategies.

## Background

Embryonal brain tumours (EBTs) are a heterogeneous, aggressive group of paediatric central nervous system (CNS) malignancies characterised by primitive cellular morphology resembling the developing foetal nervous system. They represent the most common CNS tumour group in children aged 0–4 years and account for approximately 12.2% of CNS tumours in children aged 0–14 years ^1,2^. Medulloblastoma and atypical teratoid/rhabdoid tumours (ATRT) are the most common entities within this group, accounting for 67.3% and 16.9% of EBTs, respectively ^3,4^.

Treatment of EBTs involves maximal safe resection and risk-adapted craniospinal irradiation, followed by intensive CNS-penetrant chemotherapy. For infants, radiotherapy is typically omitted or delayed until three years of age to avoid neurocognitive morbidity. As such, chemotherapy is further intensified and autologous stem-cell rescue utilised ^5,6^. These shared treatment exposures provide a rationale for considering EBTs together when evaluating treatment-related risks.

The intensity of EBT-directed therapy exposes patients to recurrent myelosuppression, mucosal barrier injury, central venous access, broad-spectrum antimicrobial use and prolonged supportive-care needs, all of which may increase susceptibility to serious infection, including IFI.

Invasive fungal infections (IFIs) are a recognised complication of intensive cancer therapy and are associated with substantial morbidity and mortality. Reported fatality rates range from 20% to 70%, depending on host, pathogen, and treatment-related factors ^7^. Frequency of IFIs varies by diagnostic group and treatment regimen. A 10-year Australian retrospective review reported risk to be highest for acute myeloid leukaemia (AML), allogeneic haematopoietic stem cell transplant (HSCT), and acute lymphoblastic leukaemia (ALL) patients, at 28.2%, 11.7%, and 10.6%, respectively ^8^. Solid tumour treatment cohorts had a lower incidence of IFIs, at 4.4% ^8^. Furthermore, discrete risk factors for IFI are recognised such as neutropaenia, corticosteroids, mucosal barrier compromise, central venous catheter use, surgical interventions, and intensive care unit (ICU) admission ^9^. Together, these exposures suggest that children receiving EBT-directed therapy may represent a distinct, under-recognised solid tumour population at risk of IFI.

The risk of IFI during intensive EBT therapy has not yet been quantified. A Brazilian study of 818 children with CNS tumours reported an overall IFI frequency of 4.6% (n=38) across all diagnoses ^10^. However, this cohort assessed all CNS tumour types, including low-grade tumours, and excluded the autologous stem-cell transplant phases of treatment. EBTs were the most common underlying diagnosis among the 38 IFI cases, accounting for 47.3% of IFIs; however, EBT-specific IFI frequency was not reported.

In this investigation, we aimed to characterise IFI during intensive EBT treatment at a single tertiary paediatric oncology centre by describing its frequency, timing and microbiology.

## Method

We retrospectively reviewed patients aged 0–18 years treated for EBTs between 2015 and 2025 at a single Australian centre. Patients were excluded if treated with non-curative intent, received less than one month of chemotherapy or if inadequate records were available (e.g. due to care at other treating centres). The clinical record was reviewed to collect diagnosis, demographics, and treatment, as well as the use of external ventricular drains (EVDs), ventriculoperitoneal (VP) shunts, stem cell support, and antifungal prophylaxis. Patients were determined to have received antifungal prophylaxis if they received a systemic antifungal agent for more than 80% of the neutropaenic phase in more than 50% of evaluable chemotherapy cycles. For patients who developed IFI, prophylaxis data was censored to onset of first IFI.

IFI diagnostic detail was collected including radiology, histopathology, and microbiology including culture, polymerase chain reaction (PCR), and galactomannan results. IFI episodes were classified as proven, probable, or possible, or modified possible, as per the EORTC/MSGERC consensus and TERIFIC study ^8,11,12^. The modified possible category was included to reflect paediatric diagnostic constraints, including limited feasibility of invasive sampling and imaging under general anaesthesia. Proven IFIs were defined as those with direct evidence of fungal invasion, including histopathological evidence of fungal elements in tissue from a normally sterile site and/or culture from a normally sterile specimen. Classification of a probable IFI required a compatible host factor, compatible clinical or radiological features, and mycological evidence, while possible IFI required a compatible host factor and clinical or radiological features without mycological confirmation. Modified possible IFI was defined in line with the TERIFIC as recognised EORTC host factors in conjunction with either chest radiographic appearances consistent with IFI when CT was unavailable, or multiple liver and/or splenic lesions suggestive of hepatosplenic fungal disease without documented candidaemia. Following initial data extraction, all suspected IFI episodes were reviewed and adjudicated by a paediatric infectious diseases physician to confirm diagnosis and classification.

IFI prevalence, clinical characteristics, and microbiology were analysed using descriptive statistics. IFI prevalence estimates were reported with 95% confidence intervals calculated using the exact binomial method. Hazard ratios were calculated using Cox proportional hazards regression model (exact partial likelihood) for sex, age, insertion of shunt or EVD, and chemotherapy protocol. Time-to-event was measured from the date of first chemotherapy to IFI symptom onset or diagnosis, whichever occurred first. Patients were censored at last follow-up or death. Statistical analyses were performed using GraphPad Prism.

## Results

Ninety-eight patients with EBTs were identified, of which 21 were excluded. 12 had insufficient records as treatment was partially delivered at other centres, and nine received less than one month of chemotherapy or were treated with non-curative intent. The final cohort included 77 patients (**Table 1**). The male:female ratio was 1.2:1 and median age was 6.4 years (range 0.2-17.4). The most frequent diagnoses were medulloblastoma (n=57; 74%) and ATRT (n=12; 16%) (**Table 2**).

**Table 1.** Clinical characteristics, diagnostic evidence and outcomes of invasive fungal infection episodes. Each row represents one IFI episode. One patient (P10) experienced two separate IFI episodes and is shown in two rows. IFI category refers to classification by modified EORTC criteria. Cycle refers to the chemotherapy cycle during which the IFI occurred. Abbreviations: Y, yes; N, no; ATRT, atypical teratoid/rhabdoid tumour; BAL, bronchoalveolar lavage; CNS, central nervous system; CSF, cerebrospinal fluid; CT, computed tomography; EBT, embryonal brain tumour; EVD, external ventricular drain; F, female; HSCT, haematopoietic stem-cell transplantation; IFI, invasive fungal infection; M, male; mEORTC, modified European Organisation for Research and Treatment of Cancer; NOS, not otherwise specified; PCR, polymerase chain reaction; VP, ventriculoperitoneal.

| IFI cat. | Sex | Protocol | Cycle IFI diagnosed | Diagnostic evidence / microbiology |
| --- | --- | --- | --- | --- |
| Proven | M | SJMB12 | 5 | Culture: *Candida parapsilosis* - blood |
| Proven | F | SJMB12 | 4 | Radiology: Multiple hypoechoic foci involving liver, lung, spleen, kidney and CNS.; Other tests: Spleen biopsy - 18S PCR detected *Candida tropicalis*. Galactomannan and PCR were negative.; Culture: *Candida tropicalis* - spleen biopsy & BAL |
| Proven | F | SJMB12 | 2 | Culture: *Candida parapsilosis* - blood |
| Proven | 1M | SJMB12 | 1 | Culture: *Kluyveromyces marxianus* - blood |
| Proven | F | SJMB12 | 4 | Radiology: No MR brain infective changes seen.; Other tests: Serum & CSF cryptococcal antigen positive; Culture: *Cryptococcus neoformans var. grubii*. - CSF |
| Probable | M | ACNS0334 | 3 | Radiology: Marked supraglottic swelling on CT with maxillary and anterior ethmoid sinus disease opacification, no bony destruction.; Other tests: Sino-nasal washing galactomannan, histopathology and PCR were negative.; Culture: *Curvularia species* & *Mucor plumbeus* - sino-nasal washings |
| Probable | M | SJMB12 | 4 | Radiology: Bilateral perivascular pulmonary nodules with surrounding ground-glass opacification on CT.; Other tests: Serum galactomannan positive on two occasions. BAL galactomannan positive, histopathology & culture negative |
| Probable | F | EuRhab> HSCT* | 12 | Radiology: Right middle and left upper lobe collapse on CT.; Other tests: Serum galactomannan positive on two occasions.; Culture: *Aspergillus fumigatus* - sputum & faeces |
| Possible | F | ACNS0334 | 3 | Radiology: Left upper lobe cavitating nodular lesion and lobar consolidation on CT.; Other tests: BAL negative galactomannan, histopathology and culture. |
| Possible | M | SJMB03 | 4 | Radiology: Segmental lobar consolidation with patchy peribronchovascular and subpleural consolidations CT.; Other tests: BAL/biopsy not done. |
| Possible | M | ACNS0333 | 1 | Radiology: Focal lobar nodularity with atypical interlobular thickening and patchy ground-glass opacification on CT.; Other tests: BAL/biopsy not done. |
| Possible | M | SJMB03 | 4 | Radiology: Nodular consolidation with surrounding ground-glass opacification on CT.; Other tests: BAL galactomannan and PCR negative.; Culture: *Candida albicans* - BAL |
| Modified possible | M | SJMB12 | 4 | Radiology: Hypodense liver lesions on CT.; Other tests: Biopsy not done. |
| Modified possible | M | ACNS0334 | 6 | Radiology: Diffuse ground-glass density with more focal dense opacification on CT.; Other tests: BAL negative galactomannan, histopathology, PCR and culture. |
| Modified possible | F | SJMB03 | 3 | Radiology: Lobar and segmental collapse with surrounding ground glass opacification on CT.; Other tests: BAL negative for galactomannan, histopathology, PCR and culture. |

**Table 2.** Baseline demographic and clinical characteristics (N = 77). ACNS0333/0334, Children’s Oncology Group protocols ACNS0333/ACNS0334; ATRT, atypical teratoid/rhabdoid tumour; CNS, central nervous system; ETMR, embryonal tumour with multilayered rosettes; EVD, external ventr icular drain; NOS, not otherwise specified; PNET, primitive neuroectodermal tumour; SJMB03/SJMB12, St Jude Children’s Research Hospital medulloblastoma protocols SJMB03/SJMB12; VP, ventriculoperitoneal.

| Characteristic | Category | n | % |
| --- | --- | --- | --- |
| **Sex** | Male | 42 | 55 |
|  | Female | 35 | 45 |
| **Age (years)** | ≤4 | 32 | 42 |
|  | 5–9 | 20 | 26 |
|  | 10–14 | 19 | 25 |
|  | ≥15 | 6 | 8 |
| **Diagnosis** | Medulloblastoma | 57 | 74 |
|  | ATRT | 12 | 16 |
|  | PNET | 2 | 3 |
|  | ETMR | 2 | 3 |
|  | Pineoblastoma | 2 | 3 |
|  | CNS Neuroblastoma | 1 | 1 |
|  | Embryonal tumour NOS | 1 | 1 |
| **Treatment protocol** | SJMB12 | 37 | 48 |
|  | ACNS0334/ACNS0333 | 28 | 36 |
|  | SJMB03 | 10 | 13 |
|  | Other | 2 | 3 |
| **Autologous stem cell rescue** | Yes | 39 | 51 |
|  | No | 38 | 49 |
| **EVD** | Yes | 49 | 64 |
|  | No | 28 | 36 |
| **VP shunt** | Yes | 27 | 35 |
|  | No | 50 | 65 |
| **Antifungal prophylaxis** | Fluconazole | 3 | 4 |
|  | None | 74 | 96 |

Patients older than three years were treated following the St Jude Children’s Research Hospital’s SJMB12 or SJMB03 protocols (48% and 13%, respectively) whilst patients younger than three years received radiation-sparing therapy according to ACNS0334 (36%). There were two exceptions (3%) based on the individual case. 39 patients (51%) received autologous stem cell rescue. All patients treated on ACNS0334 and SJMB03 received stem cell rescue, as did one other patient. An EVD was inserted in 49 patients (64%) and a VP shunt in 27 patients (35%). Three patients (4%) received antifungal prophylaxis, all with fluconazole (Table 2). No patients with an IFI received antifungal prophylaxis.

Fourteen patients experienced 15 IFI episodes giving a patient-level prevalence of any IFI of 18.2% (95% CI, 10.3–28.6%). There were five proven IFIs, three probable, four possible and three modified possible (Table 1). Seven patients had a proven or probable IFI, giving a prevalence of 9.1% (95% CI, 3.7%–17.8%). IFI occurred during chemotherapy phase of treatment for all patients with 73% of IFIs occurring during cycle 4 or beyond. Median time from chemotherapy commencing to IFI diagnosis was similar between patients treated with SJMB03/SJMB12 (97.5 days, IQR 78.75–121.25) and those treated with radiation-sparing ACNS0334/ACNS0333 protocols (98.5 days, IQR 51.75–160.25).

Nine IFI episodes had mycological pathogen evidence, although only five episodes (55.6%) involved organism recovery from a normally sterile site. *Candida* species were cultured in four (44%), non-*Candida* yeasts in two (22%), *Aspergillus fumigatus* in one (11%), and non-*Aspergillus* moulds in two (22%). One further probable pulmonary aspergillosis episode was supported by serial positive serum galactomannan and positive bronchoalveolar lavage (BAL) galactomannan, despite negative histopathology and culture (Table 1). The most frequent sites of infection were pulmonary (40%) and blood (20%). For two patients, IFI resulted in a chemotherapy delay of more than one month. There were no deaths directly attributed to IFI.

Exploratory Cox proportional hazards analyses did not identify clear associations between sex, age at diagnosis, EVD, or VP shunt (**Table 3**). Statistical significance was defined as a two-sided p-value <0.05. In analyses restricted to proven/probable IFI, ACNS0334/ACNS0333 was associated with a lower estimated hazard compared to SJMB12 (HR 0.062; 95% CI, 0.002– 0.78; p=0.031). Each additional year of age was associated with a lower estimated hazard, although this did not reach statistical significance (HR per year increase, 0.81; 95% CI, 0.56– 1.03; p=0.089).

**Table 3.**
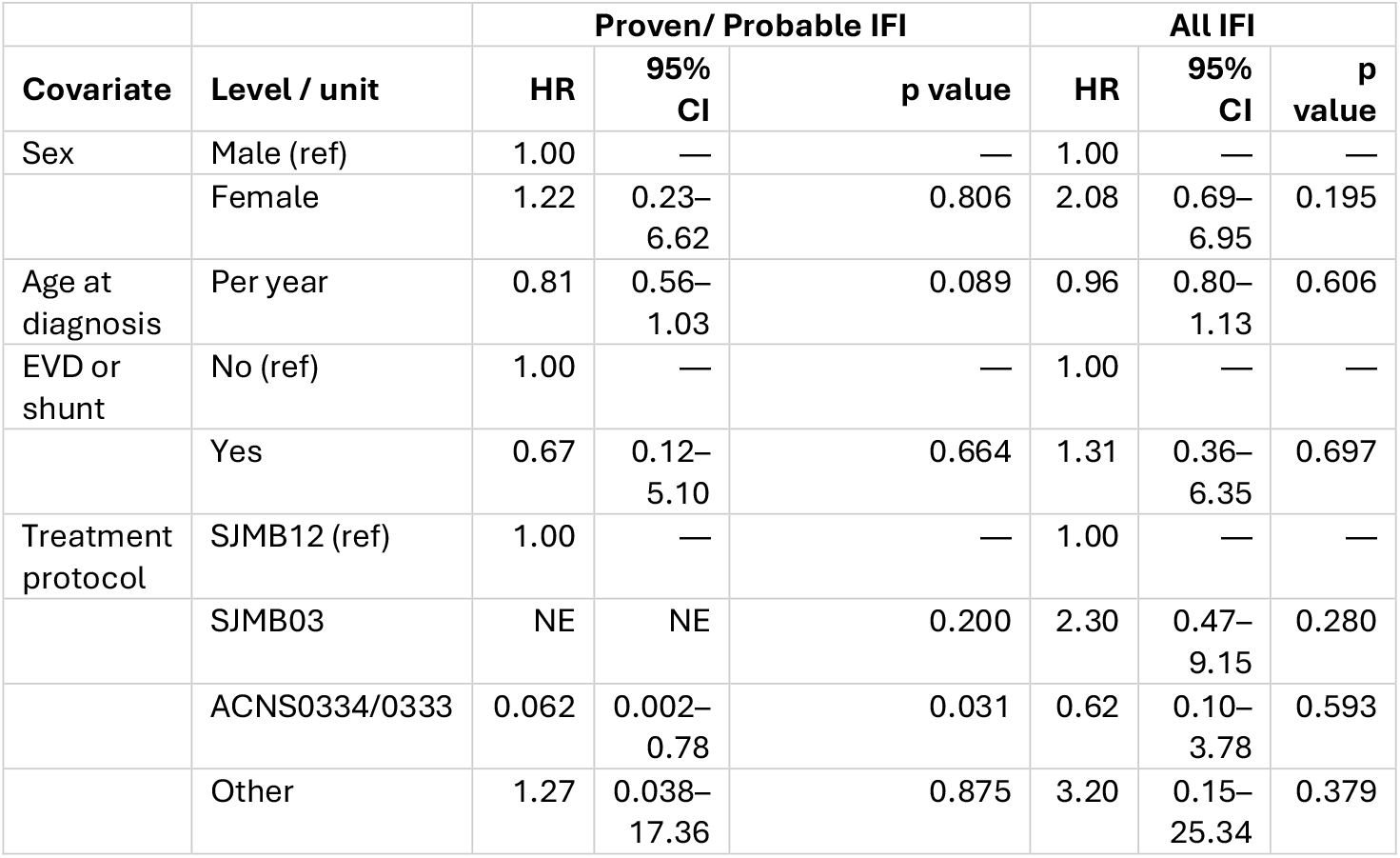
Multivariable Cox proportional hazards model for time to IFI (N = 77). NE = not estimable. Time scale: days. Time-to-event analyses were performed at the patient level using time to first qualifying IFI. Event counts were 14 patients for any IFI and 7 patients for proven/probable IFI. One patient experienced two IFI episodes and was counted once in each analysis.

## Discussion

In this single-centre cohort of children receiving intensive treatment for EBTs, IFI occurred in 18.2% of patients overall, with proven/probable IFI in 9.1%. This frequency is higher than expected for paediatric solid tumour cohorts and approaches rates reported in traditionally higher-risk paediatric oncology groups, such as children treated for acute haematological malignancies.

For comparison, the Australasian TERIFIC study reported prevalences in children of 28.2% in AML, 11.7% in allogeneic HSCT, 10.6% in ALL and 4.4% in solid malignancies using the same IFI classification criteria ^8^. Although inclusion of modified possible IFI may increase the overall estimate, the burden remained notable when restricted to proven/probable IFI, which occurred in 9.1% of patients. A Brazilian cohort of children with CNS tumours reported an overall IFI frequency of 4.6%, although this included all CNS tumour types, excluded autologous stem-cell transplant phases, and did not report EBT-specific incidence ^10^. To the best of our knowledge, ours is the first study to report IFI risk in a denominator-defined cohort of children receiving intensive EBT-directed therapy. Our findings suggest that children receiving EBT-directed therapy may represent a distinct subgroup within paediatric solid tumour care.

Non-mould infections were more common than mould infections, accounting for 6/9 episodes (67%) of identified pathogens overall. This microbiological pattern appears more similar to reports in paediatric solid tumour cohorts, where yeast/non-mould infections predominate, than to AML, ALL or HSCT cohorts in which mould infections are relatively more frequent. In the Australian TERIFIC cohort, yeasts accounted for 77.4% of identified pathogens in solid tumours and 88.9% in neuroblastoma, compared with 43.9% in ALL, 38.9% in AML and 46.7% in HSCT ^8^. This may reflect a pattern in our cohort of IFI risk related to barrier injury and device-associated bloodstream infection, as proposed in other solid tumour cohorts, rather than the prolonged profound neutropaenia and inhalational mould exposure that more typically drive IFI risk in haematology and HSCT populations. The small number of microbiologically identified episodes in our cohort limits interpretation however.

The only factor associated with a lower hazard of proven/probable IFI was treatment with ACNS0334/0333 protocols. This association may reflect protocol-specific differences in treatment exposure, including the timing and intensity of induction chemotherapy, mucosal toxicity, duration of neutropenia, and rapid count recovery following autologous stem-cell rescue.

In our cohort, only three children (4%) received antifungal prophylaxis, in keeping with national and international guidelines which do not specifically identify children receiving EBT therapy as a group requiring prophylaxis. This is in contrast to other groups such as children with AML and those receiving allogeneic HSCT, where antifungal prophylaxis is recommended during periods of highest vulnerability ^13^. While this retrospective study cannot determine whether prophylaxis would reduce IFI in this population, the frequency of IFI, including proven/probable IFI, supports prospective evaluation of risk-adapted antifungal prophylaxis and provides a clinical rationale for centres to review prevention strategies in children receiving intensive EBT therapy.

This study has limitations. Its retrospective single-centre design limits generalisability and introduces potential ascertainment bias. The small number of IFI events limited statistical power, particularly for proven/probable IFI, and prevented robust multivariable modelling of correlated treatment exposures such as age, protocol, and autologous stem-cell rescue. Finally, the microbiology observed in this single-centre Australian cohort may reflect local environmental exposures, climate, hospital infrastructure, and antifungal practices, limiting generalisability to centres with different endemic fungi or supportive-care protocols.

## Conclusion

Intensive EBT therapy was associated with a substantial burden of IFI at a frequency higher than expected for paediatric solid tumour cohorts. IFI occurrence was associated with treatment disruption and prolonged morbidity. These findings support recognition of children receiving EBT-directed therapy as a distinct IFI-risk group within paediatric oncology and that centre specific consideration of antifungal prophylaxis is warranted.

**Figure 1.**
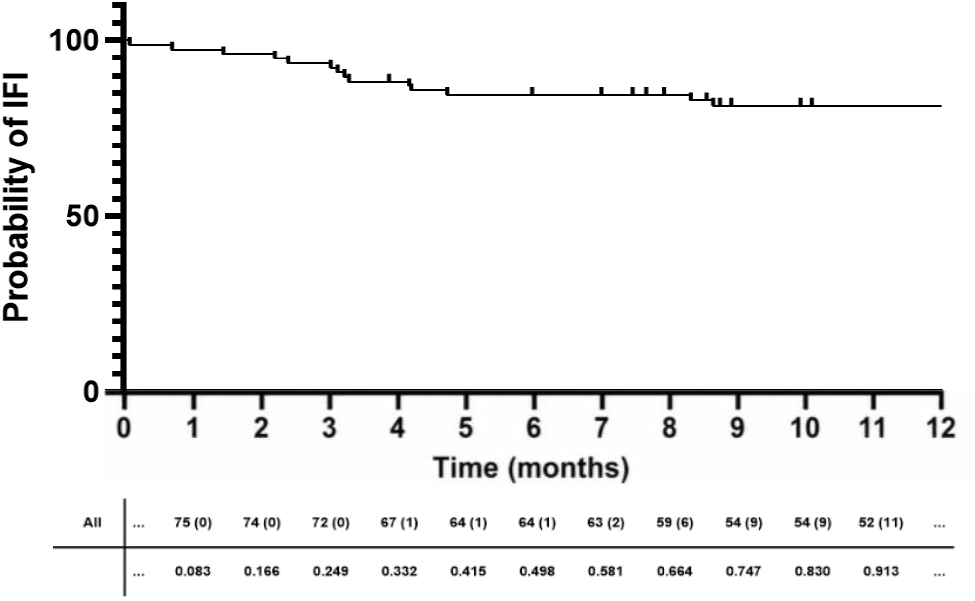
Time from commencement of chemotherapy to first invasive fungal infection. Kaplan–Meier curve showing IFI-free survival from the date of first chemotherapy. Patients were censored at last follow-up or death if no IFI occurred. Time is shown in months.

## Data Availability

All data produced in the present study are available upon reasonable request to the authors

## Required Statements

### Ethics

This minimal-risk retrospective study received ethical review and governance authorisation through the Melbourne Children’s Campus, incorporating The Royal Children’s Hospital, Murdoch Children’s Research Institute and the University of Melbourne Department of Paediatrics (MCC DERP reference 3771). Initial approval was granted on 28 March 2024, with amendment approval on 29 September 2025. The requirement for individual informed consent was waived because the study involved retrospective review of existing clinical data.

### Authorship

Study conception and design: S.M.C., M.C., and G.M.H. Data collection: S.M.C., A.C., and G.M.H. Data analysis: S.M.C., D.A.K.Q., D.D.E., and G.M.H. Statistical analysis: S.M.C. and G.M.H. Data interpretation: S.M.C. and G.M.H. Manuscript drafting: S.M.C., A.C., and G.M.H. Critical revision of the manuscript: all authors. Study supervision: G.M.H. Project administration: S.M.C. All authors approved the final manuscript.

### Data Availability

The data underlying this article will be shared on reasonable request to the corresponding author.

